# A seq2seq model to forecast the COVID-19 cases, deaths and reproductive *R* numbers in US counties

**DOI:** 10.1101/2021.04.14.21255507

**Authors:** Yanli Zhang-James, Jonathan Hess, Asif Salekin, Dongliang Wang, Samuel Chen, Peter Winkelstein, Christopher P Morley, Stephen V Faraone

## Abstract

The global pandemic of coronavirus disease 2019 (COVID-19) has killed almost two million people worldwide and over 400 thousand in the United States (US). As the pandemic evolves, informed policy-making and strategic resource allocation relies on accurate forecasts. To predict the spread of the virus within US counties, we curated an array of county-level demographic and COVID-19-relevant health risk factors. In combination with the county-level case and death numbers curated by John Hopkins university, we developed a forecasting model using deep learning (DL). We implemented an autoencoder-based Seq2Seq model with gated recurrent units (GRUs) in the deep recurrent layers. We trained the model to predict future incident cases, deaths and the reproductive number, *R*. For most counties, it makes accurate predictions of new incident cases, deaths and R values, up to 30 days in the future. Our framework can also be used to predict other targets that are useful indices for policymaking, for example hospitalization or the occupancy of intensive care units. Our DL framework is publicly available on GitHub and can be adapted for other indices of the COVID-19 spread. We hope that our forecasts and model can help local governments in the continued fight against COVID-19.

## Introduction

The global pandemic of COVID-19, first identified from an outbreak in Wuhan, China at the end of December 2019, has drastically changed our societies and economies all over the world. As of the beginning of 2021, the virus has infected more than 86 million people and claimed almost two million lives across the globe. The US has reported approximately 24% of the world’s total cases and 19% of the world’s total deaths and has been reporting the highest numbers of daily new infections across the world since November 2019. The cooler weather, the reopening of schools and businesses in the fall, holiday travel and gatherings, and the evolving infectiousness of the virus have contributed the rapidly rising cases in many regions.

Accurate forecasting models are essential for government policymaking during the fight against the virus. A variety of methodologies have been used to forecast the spread of COVID-19. Most have used traditional epidemiological models such as the susceptible-infected-removed (SIR), or Susceptible-Exposed-Infectious-Removed (SEIR) models. These models use ordinary differential equations and rely on assumptions such as the transmission characteristics of the virus. Machine learning and deep learning methods have also been used. They provide a complementary, data-driven approach that does not rely on epidemiological assumptions. The most important prerequisite for deep learning models is access to a large amount of data from which to learn. While deep learning models were limited by the availability of the data in the early months, the long-stretch of the pandemic worldwide has, unfortunately, provided ample data for more accurate modeling and longer-term forecast.

Many teams and methods have been used to model and forecast the spread of the pandemic. Currently, over 50 teams from the globe are participating in the COVID-19 Forecast Hub (www.covid19forecasthub.org), led by the Reich Lab at the University of Massachusetts. The Forecast Hub coordinates with the US Centers for Disease Control and Preventions (CDC) to generate weekly forecasts of cases, deaths, and hospitalizations in the US for up to four weeks in the future. The ensemble forecast, which uses the median prediction across a few dozen of eligible models, are published weekly on the CDC website (Ray, Wattanachit et al. 2020). Most of the forecasts focus on national and state outcomes. Only a few teams have generated forecasts for US counties. The weekly published ensemble models only forecast incident cases at the county-level. They do not forecast deaths or other outcomes.

The main goal of this study was to improve the accuracy of county-level forecasts and to increase the types of outcomes forecast at the county level to include R values and deaths. Another strength of our model is the inclusion of multimodal predictors. We curated an array of relevant demographic indicators such as population density, rural/urban ratios, and a number of chronic health risk factors that have been linked to the risk of more severe COVID-19 infections. We demonstrate that our model learned patterns of input features to make accurate predictions of new incident cases, deaths, and R values up to 30 days in the future. Our framework can also be used to predict other targets that are useful indices for policymaking, for example hospitalization or the occupancy of intensive care units (ICU).

## Methods

### Data source

The time series for daily cumulative cases and deaths were downloaded from the COVID-19 Data Repository by the Center for Systems Science and Engineering (CSSE) at Johns Hopkins University (Dong, Du et al. 2020). Incident cases and deaths were generated from the cumulative series and seven-day moving averages (7-MA) were used as input sequences for the prediction models.

We computed the daily reproduction number, *R*, time series using a sliding weekly window for each county based on the methods described in Cori, et al (2013) and implemented in the R Statistical Software package EpiEstim. Negative numbers in the incident cases series due to reporting errors were set to 0. *R* indicates how fast the virus is spreading by computing the average number of secondary cases of disease caused by a single infected individual during the whole infectious period. Higher *R* values that are above 1 indicate the increasing spread of the virus in the community. The goal of epidemic control is to bring the *R* values to below 1 (as close to 0 as possible).

Google has released data on daily mobility score changes reflecting the effects of social distancing and local lockdown measures, as well as school and business status. The Google Community Mobility Reports (https://www.google.com/covid19/mobility/) contain percentages of movement changes over six types of communities: retail & recreation, grocery & pharmacy, parks, transit, workplaces, residential regions. The changes were based on “baseline” values reported from January – February 2020. We computed a single daily mobility score by averaging the percentage changes from five categories (retail & recreation, grocery & pharmacy, parks, transit, workplaces) and the negative values of residential changes. All six categories showed highly significant correlations and are also significantly correlated with the mean daily mobility score. The daily mean mobility scores were missing in 24% of the days, for which we used the mean daily mobility scores of the previous scores. Mobility scores prior to February 15, 2020 were considered to be baseline, therefore set to 0%.

Population size, population density, rural/urban ratio, and other demographic information such as sex, age and racial composition, were obtained from the 2019 National and State Population Estimates from the US Census Bureau. The numbers of ICU beds (per 1,000 population) were obtained from the 2019 Kaiser Family Foundation Survey (KFF). Other health risk factors, such as differential use of preventive services (preventable hospitalization rate, insurance coverage, mammograph and flu vaccine immunization rates), chronic health conditions (diabetics, smoking and obesity), socioeconomic factors (household income, poverty, education, housing density, unemployment rate), as well as an air quality index, were obtained from the Behavioral Risk Factor Surveillance System (BRFSS). A total of 28 county-level variables are included as base-line features. All data were linked by counties’ 5-digit FIPS (Federal Information Processing Standards) codes.

### Seq2Seq Model

Our prediction model used the Sequence-to-sequence learning framework (Seq2Seq) that powers language-processing applications like Google Translate (Sutskever et al., 2014). The Seq2Seq algorithm trains a model to convert sequences from one domain (e.g. past COVID-19 cases and deaths, resident mobility, or infection numbers, etc.) to sequences in another domain (e.g. future COVID-19 cases). Seq2Seq generation is flexible in that input sequences and output sequences can have different lengths. However, the information on the entire input sequence is required in order to predict the target sequence.

Seq2Seq uses feed-forward recurrent neural networks (RNNs) that are specialized for mapping sequences. Our Seq2Seq model (shown in Figure 1), uses Gated Recurrent Units (GRUs, Dey and Salem 2017) in the RNN layers. The model is consisting of three parts: an encoder, an encoding vector (generated from the input sequence), and a decoder (Cho et al., 2014; Sutskever et al., 2014). Our Seq2Seq model takes ‘m’ days data as input and predicts COVID-19 cases for ‘n’ future days. The encoder generates a comprehensive aggregated representation of the input ‘m’ days of the sequence. Our encoder implementation is a stack of ‘m’ GRU units, where each unit accepts input features (i.e., COVID-19 daily cases, mobility and deaths, etc.) from a single day of the input sequence, aggregate the information, and propagates the aggregated information forward *via* hidden state h_i_ to the next GRU unit in the sequence. The final hidden state produced from the encoder represents the encoding of the entire input sequence. This vector aims to encapsulate the information for all input elements in order to help the decoder make accurate predictions. The generated encoding is provided as the initial hidden state input of the decoder. That means, h_m+1_=s_1_ in the Figure 1. The decoder predicts the output sequence taking ‘Encoding of the input sequence’ as input. It is a stack of ‘n’ GRU units where each unit (*j*th unit) predicts an output/prediction of COVID-19 cases (for the *j*th future day). Each GRU unit accepts a hidden state from the previous unit and produces an output as well as its own hidden state.

**Figure 1.**
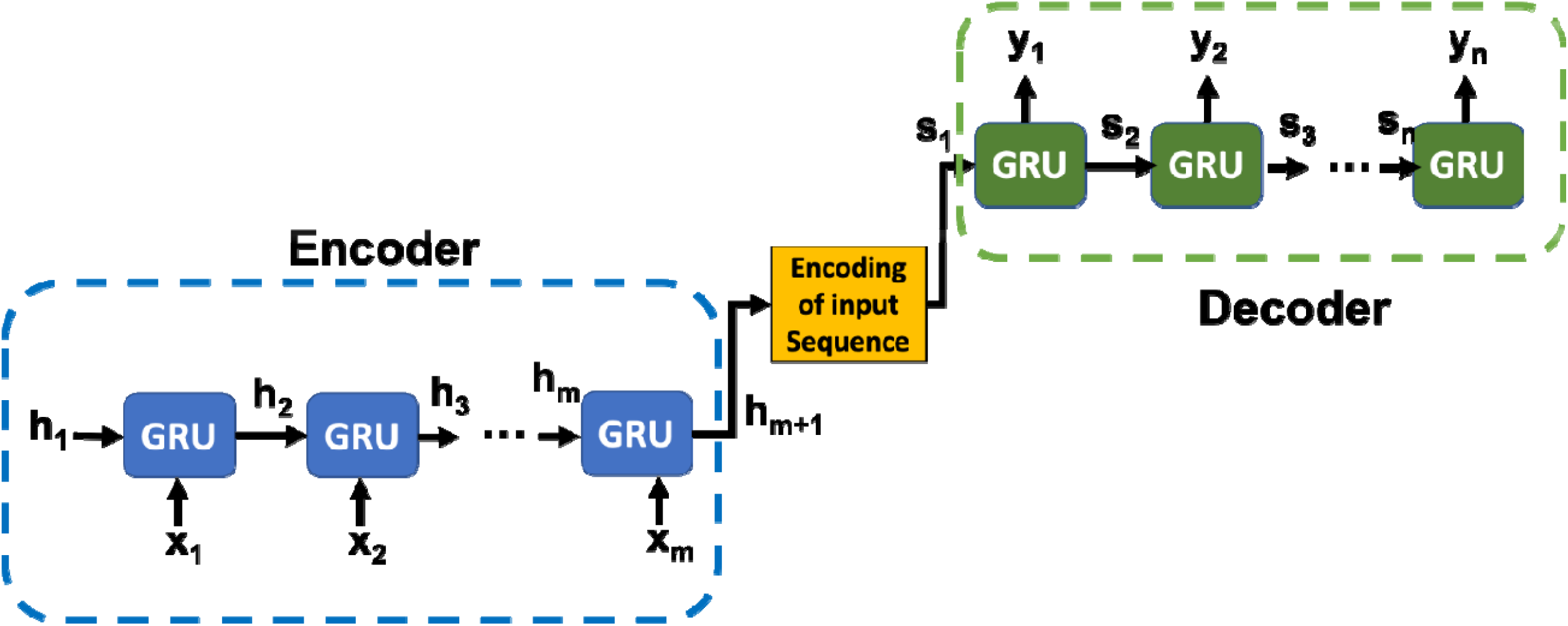
Sequence-to-sequence learning framework1

The power of this model lies in the fact that it can map sequences of different lengths to each other. The *i*th inputs and *j*th outputs are not necessarily correlated and the sequence lengths can differ.

### Model selection and hyperparameter tuning

Hyperparameter tuning for the Seq2Seq model was carried out using the Keras API (version 2.3.1), the TensorFlow library (version 1.14.0) and HyperOpt (Bergstra, Yamins et al. 2013). The encoder and decoder have two layers. Our search space included layer sizes ranging from three to 5000. We used the Adam optimizer and tuned the learning rate and decay in the range of 1*e*-6∼1*e*-2. We tuned the L2 regularization parameter in the range 1*e*-7 – 1*e*-3. L2 regularization was applied to both the weights and biases. We tuned the mini-batch size in the range 4 – 256. Early stopping was used to avoid overfitting. Models were trained using the mean square error (MSE) as the loss function. The best model hyperparameters were chosen based on the lowest validation loss. We inspected both the training and validation losses to ensure that models were not overfitting.

Learning and remembering a long input sequence can be challenging for a recurrent network. Short input sequences, on the other hand, may not provide enough information for learning patterns and making accurate forecasts. To determine the optimum input sequences, we used HyperOpt to search the above hyperparameter spaces for three input sequence sizes: 60, 90 and 120 days of input data (cases, deaths, Google mobility, and *R* values). The model predicted outcomes for each day over the next 30 days. The optimum input sequence length was determined as the one that resulted in models with the lowest validation losses.

### Model training and forecast evaluation

From the 3242 counties (or county-equivalents) in the US states and territories, 2780 had data on the input features we selected for modelling. We randomly split these counties into training (n=1983, 71.3%), validation (n=398, 14.3%) and test sets (n=399, 14.4%). We used the training and validation sets for the hyperparameter search and for learning the model weights and biases. As Supplementary Figure 1 shows, the input sequences were generated from the whole series of data on a N-day sliding window letting N be the best length of input days. For each county, day 1 was determined as the first day when the first case was reported in the county. Models were trained on stacks of input sequences with the fixed optimum length N to predict the next 30 days of outcomes. Training stopped when the validation MSE did not decrease over two consecutive epochs. We evaluated model performance on the test set using data up to November 30, 2020. Accuracy metrics include the relative error (RE= (predicted value-observed value)/observed value), and the interclass correlation coefficient between the predicted and the observed values using a two-way random effect model (ICC(2,1)). In addition to examining the prediction accuracy for the test set on the days that we had data from the training and validation set to train the model, we also examined the model’s performance as a historical evaluation by comparing the forecasted values at a given date with the actual values that were reported later. For the historical performance, we reported Pearson’s correlation coefficient (Pearson’s *r*) and RE for the November 30 forecast to evaluate how well the model predicted future cases or deaths over the next 30 days.

Finally, using our 30-day forecast, we generated the 4-week ahead weekly forecast (total weekly incident cases) that we submited to the COVID-19 Forecast Hub for inclusion in the CDC ensemble model. We compared the historical performance of our forecast with that of the CDC ensemble model as well as the models of the individual participating teams on two forecasting dates, November 23 and 30, 2020. In addition to the ICC(2,1) correlation coefficients between each team’s forecasted values with the actual reported values and the REs, we also reported the absolute relative errors (ARE= absolute(predicted value-observed value)/observed value), the median of which represented the overall degree of deviation of predicted values from the observed values better than the median of REs.

## Results

### Determine the optimum input sequence length

The initial hyperparameter space search for models using various length of input sequences showed that the lowest training and validation MSEs were achieved by using 90 days of input sequences (Supplementary Figure 2). Using either shorter (60 days) or longer (120 days) input resulted in higher training and validation MSEs. Therefore, we built overlapping sliding windows of 90 days from the input sequences to train the model for predicting the next 30 days as illustrated in Supplementary Figure 1.

### Test set evaluation

Model predicted incident cases for subsequent 30-day periods were in highly significant agreement with the seven day moving average for the actual reported cases (Figure 2A, ICC(2,1) = 0.47, 95%CI: 0.43-0.50, F=36.83, p<0.0001). Model predicted incident deaths were also in highly significant agreement with the seven-day moving average of the actual reported deaths (Figure 2B, ICC(2,1) = 0.32, 95%CI: 0.29-0.36, F=10.36, p<0.0001). Figure 2B highlights several outliers (as red dots). These were due to an irregularity of death reporting in Passaic county, New Jersey on June 25, 2020 (Figure 2C). Excluding Passaic county, ICC(2,1) was 0.38 (95%CI: 0.34 - 0.43, F=14.6, p<0.0001). The R values predicted from our model were also highly significantly correlated with the R values calculated from the actual cases (ICC(2,1) = 0.57, 95%CI: 0.53-0.61, F=13.37, p<0.0001, Figure 2D).

**Figure 2.**
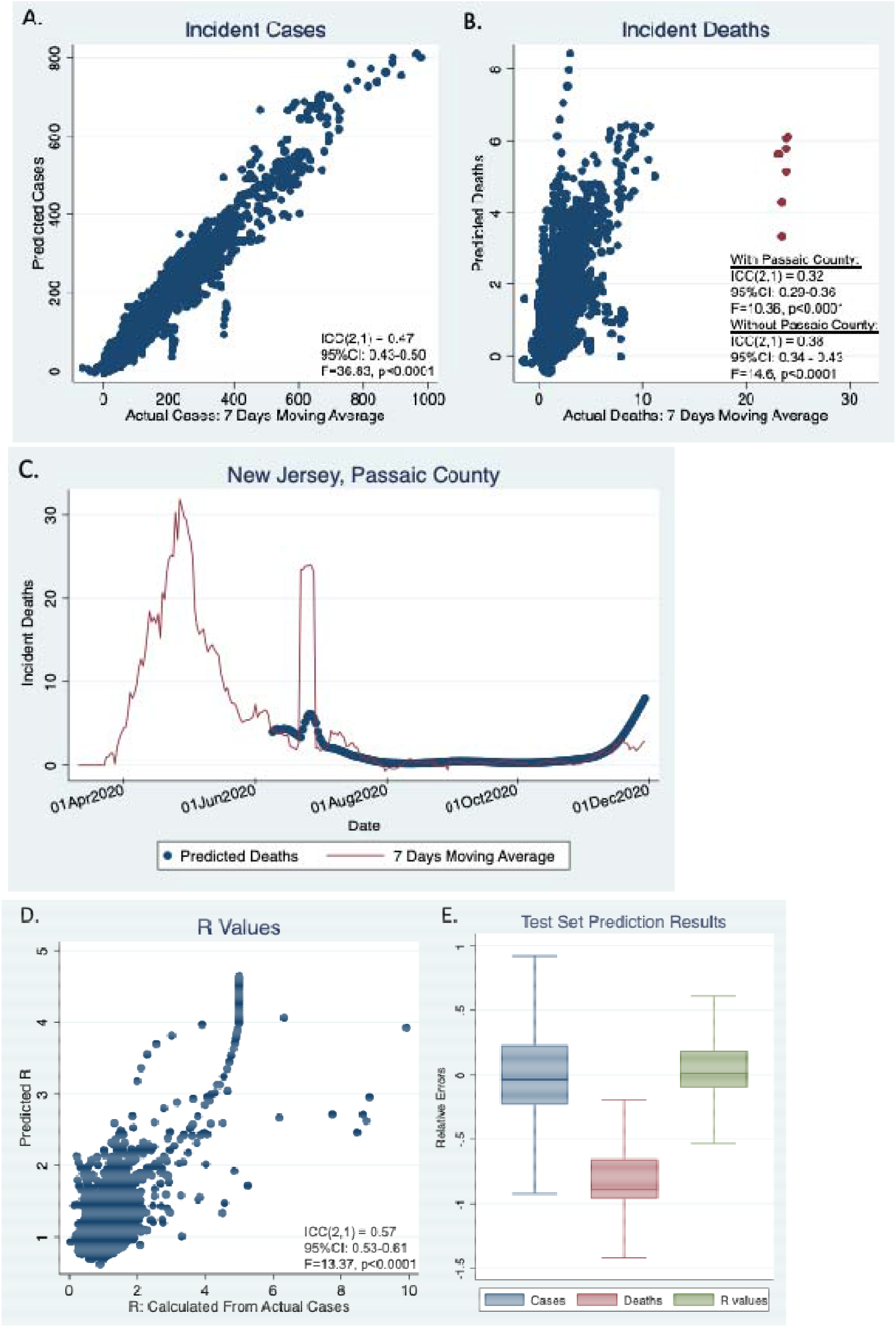
Test set results of cases and deaths.

The relative errors (REs) are plotted in Figure 2E. The median REs are close to 0 for both incident cases (−0.04) and R values (0.01), despite that statistically they are significantly different from 0 (p<.0001) due to the large sample sizes. On the contrary, the median RE is - 0.89 for incident deaths, and RE distribution is significantly deviated from 0, indicating that our model is significantly under-predicting deaths.

### Forecast evaluation

The performance of the model was evaluated using the later reported cases and deaths. Using the forecasts generated on November 30 as an example, we show in Supplementary 3A that the daily predicted cases from all counties for the future 30 days were significantly correlated with the seven-day moving average of actual reported cases. Over the 30 days, Pearson’s *r* declined slowly but remained highly significant, with the highest correlation of *r* = .98 found for the first day to ∼.70 for the furthest forecast days (all p<.001). Supplementary Figure 3B shows the predicted deaths vs the actual deaths. The daily correlation *r* ranged from 0.87 to 0.63 (all p<.001). Supplementary Figure 3C shows the predicted R values versus the R calculated from the actual cases. The daily correlation r ranged from 0.64 - 0.29 (all p<.001).

Examination of the RE distributions (Supplementary Figure 3D) shows accurate case forecasts based on their consistent and small RE values that are close to 0 (median RE −0.06). Deaths forecasts, although having a smaller RE quantile range, consistently deviated from 0 in the negative direction (median RE −0.76). The R values have very small quantile ranges and an overall median RE (−0.008) close to zero.

Similar results were found for forecasts generated on other dates (not reported). The 30-day ahead forecasts is updated weekly on Mondays and is archived in our GitHub repository (https://ylzhang29.github.io/UpstateSU-GRU-Covid/).

### Comparison with other models

We compared our model forecast results at two time points, November 23 and November 30, with the corresponding results from the CDC ensemble model and individual models from the participating teams. Twelve teams and their models have reported forecasts for US counties, including the CDC ensemble model. Only case forecasts have been reported from these teams. Note that our forecasts at these two dates were not submitted and included in the ensemble model. The total numbers of counties forecasted by each team/model and their median REs are summarized in Supplementary Table 1.

To compare the models on the forecasts of the same set of counties, we excluded three teams that reported < 50% of the total counties. Nine teams/models, including the ensemble models were compared with our model using only the common set of counties that all models reported forecasts on. Figure 3A plots the AREs distributions of the two forecasts on each of the four weeks ahead from these nine models and our model. Figure 3B plots the overall median AREs for each team over the four weeks ahead forecast. Table 1 lists additional evaluation metrics including both median RE and median ARE. The Wilcoxon rank-sum test was used to compare our model’s ARE with other models’ ARE (see Table 1). Table 2 shows the ICC(2,1) between the predicated and actual cases for each models’ forecast. Overall, these results showed that while all models were able to produce accurate forecasts that were in significant agreements with the actual observed values, there are still large prediction uncertainties in many counties, evidenced by the large RE and ARE medians (deviation from 0). The prediction uncertainties increase as the forecast interval increases (Figure 3B).

**Figure 3.**
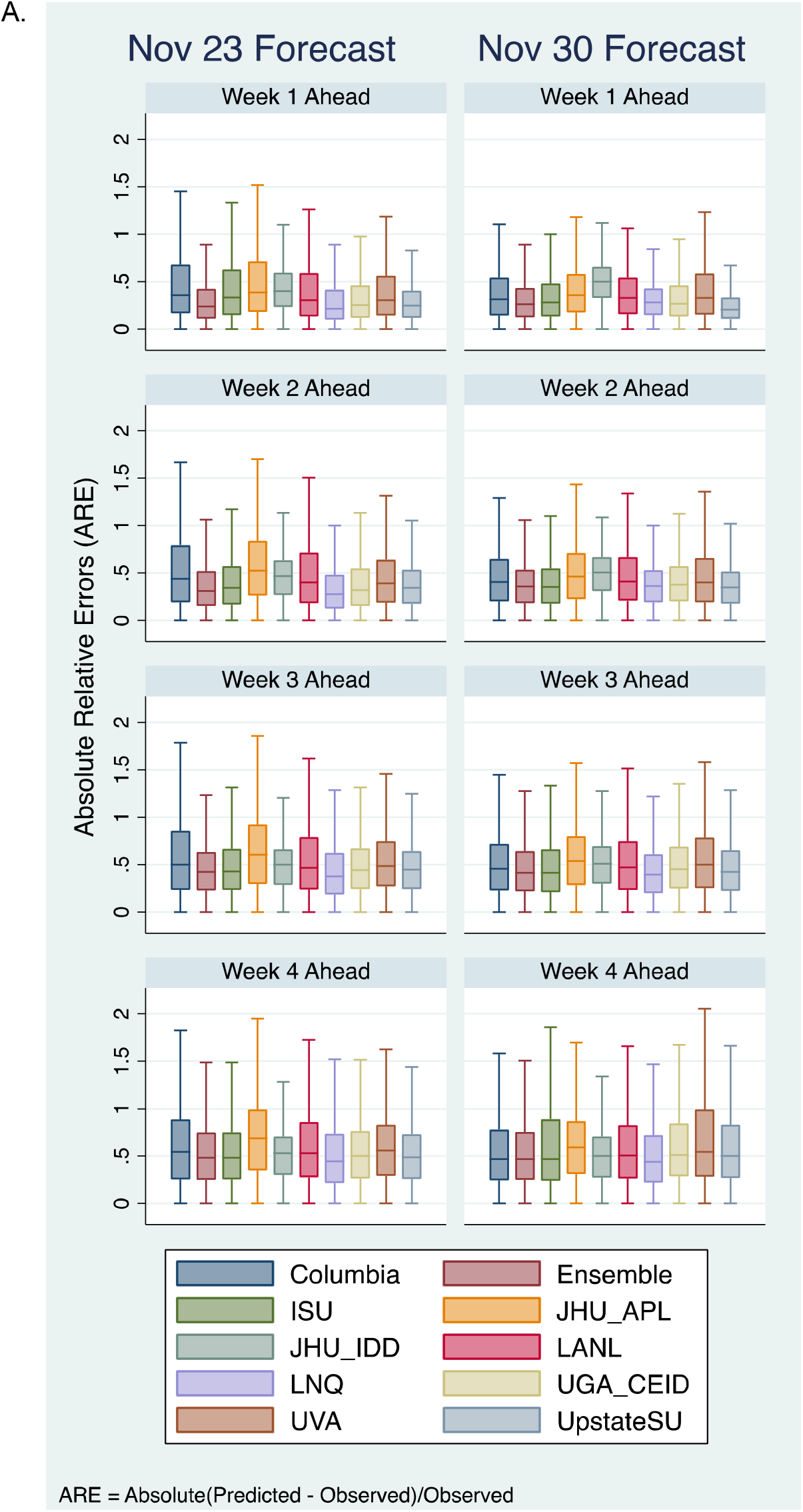
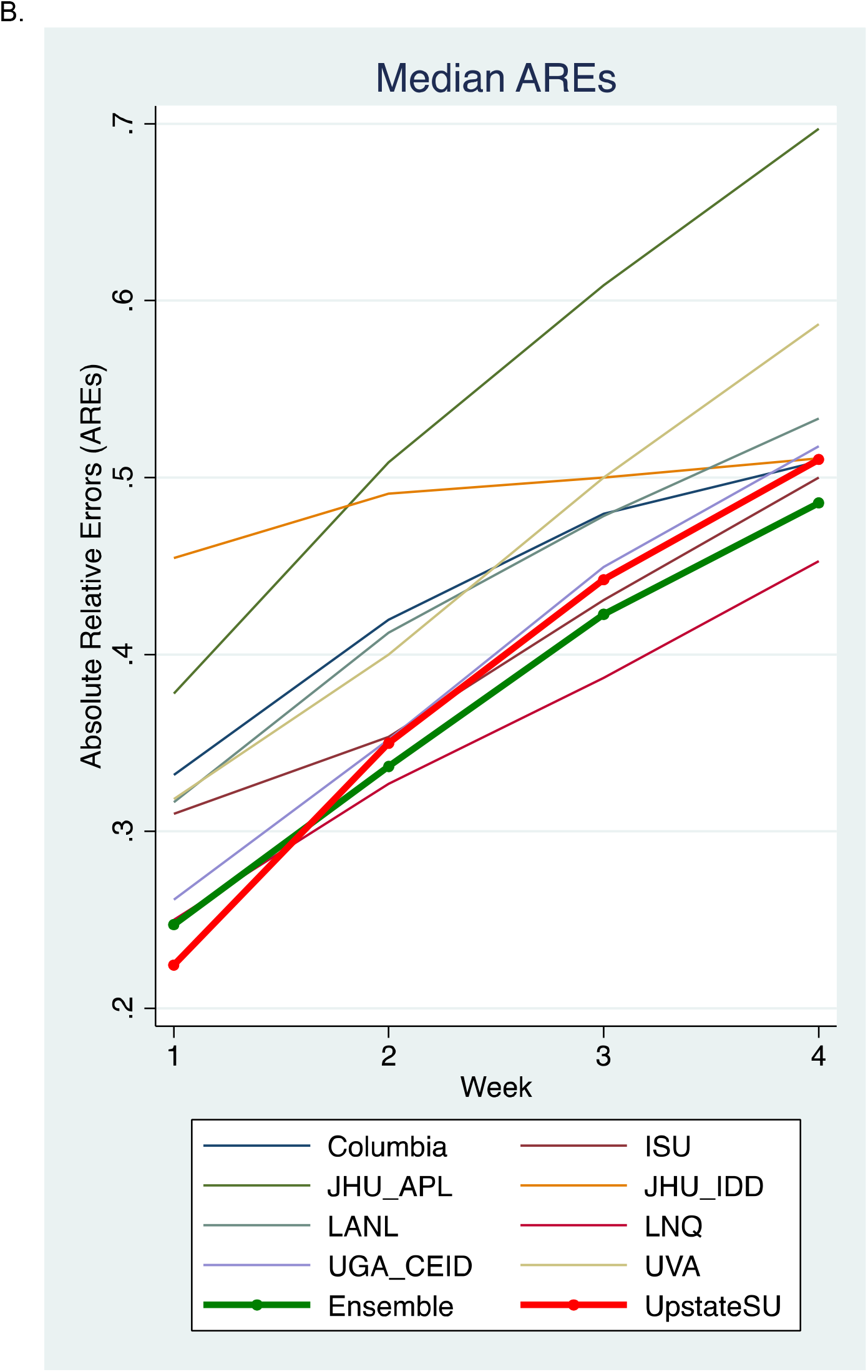
A. Comparison of ARE distribution of our model with nice additional models that reported to CDC, including the ensemble model. B. Median AREs for each team/model were plotted over four-week ahead forecasts. *For model names and team information, see CDC’s COVID-19 forecast model descriptions: https://github.com/cdcepi/COVID-19-Forecasts/blob/master/COVID-19_Forecast_Model_Descriptions.md

**Table 1.** Comparisons of county-level case forecast from different teams and models. Total numbers of counties that are present in all models are listed for each week’s forecast. RE, median relative errors. ARE, median absolute relative error.

| Nov 23 Forecast |  |  |  |  |  |  |  |  |  |  |  |  |
| --- | --- | --- | --- | --- | --- | --- | --- | --- | --- | --- | --- | --- |
| Team | Week 1<br>(Counties in all models N=2737) |  |  | Week 2<br>(Counties in all models N=2740) |  |  | Week 3<br>(Counties = in all models N=2738) |  |  | Week 4<br>(Counties in all models N=2736) |  |  |
|  | RE<br>(Median) | ARE<br>(Median) | ARE differ from<br>UpstateSU | RE<br>(Median) | ARE<br>(Median) | ARE differ from<br>UpstateSU | RE<br>(Median) | ARE<br>(Median) | ARE differ from<br>UpstateSU | RE<br>(Median) | ARE<br>(Median) | ARE differ from<br>UpstateSU |
| Columbia | -0.02 | 0.36 | p<.0001 | -0.15 | 0.44 | p<.0001 | -0.21 | 0.50 | p<.0001 | -0.23 | 0.55 | p<.0001 |
| Ensemble | 0.02 | 0.24 | p=.070 | -0.13 | 0.31 | p<.0001 | -0.19 | 0.42 | p=.006 | -0.15 | 0.49 | p=.387 |
| ISU | 0.17 | 0.34 | p<.0001 | -0.02 | 0.35 | p=.829 | -0.11 | 0.44 | p=.195 | -0.08 | 0.50 | p=.871 |
| JHU_APL | 0.01 | 0.39 | p<.0001 | -0.06 | 0.55 | p<.0001 | -0.06 | 0.66 | p<.0001 | 0.06 | 0.76 | p<.0001 |
| JHU_IDD | -0.38 | 0.40 | p<.0001 | -0.45 | 0.47 | p<.0001 | -0.46 | 0.50 | p<.0001 | -0.47 | 0.53 | p=.008 |
| LANL | 0.06 | 0.30 | p<.0001 | 0.00 | 0.41 | p<.0001 | -0.05 | 0.48 | p=.066 | -0.08 | 0.55 | p<.0001 |
| LNQ | 0.05 | 0.22 | p<.0001 | -0.05 | 0.28 | p<.0001 | -0.04 | 0.38 | p<.0001 | 0.04 | 0.45 | p=.001 |
| UGA_CEID | 0.06 | 0.25 | p=.317 | -0.11 | 0.32 | p=.003 | -0.18 | 0.44 | p=.417 | -0.15 | 0.51 | p=.552 |
| UpstateSU | -0.09 | 0.25 | NA | -0.21 | 0.35 | NA | -0.21 | 0.45 | NA | -0.11 | 0.50 | NA |
| UVA | 0.07 | 0.31 | p<.0001 | -0.10 | 0.40 | p<.0001 | -0.14 | 0.50 | p<.0001 | -0.10 | 0.58 | p<.0001 |
| Nov 30 Forecast |  |  |  |  |  |  |  |  |  |  |  |  |
|  | Week 1<br>(Counties in all models N=2764) |  |  | Week 2<br>(Counties in all models N=2762) |  |  | Week 3<br>(Counties in all models N=2761) |  |  | Week 4<br>(Counties in all models N=2750) |  |  |
|  | RE<br>(Median) | ARE<br>(Median) | ARE differ from<br>UpstateSU | RE<br>(Median) | ARE<br>(Median) | ARE differ from<br>UpstateSU | RE<br>(Median) | ARE<br>(Median) | ARE differ from<br>UpstateSU | RE<br>(Median) | ARE<br>(Median) | ARE differ from<br>UpstateSU |
| Columbia | -0.17 | 0.31 | p<.0001 | -0.27 | 0.41 | p<.0001 | -0.25 | 0.46 | p=.019 | -0.13 | 0.48 | p<.0001 |
| Ensemble | -0.18 | 0.26 | p<.0001 | -0.24 | 0.36 | p=.282 | -0.21 | 0.42 | p=.319 | -0.05 | 0.48 | p<.0001 |
| ISU | 0.00 | 0.29 | p<.0001 | -0.07 | 0.35 | p=.788 | -0.04 | 0.42 | p=.536 | 0.13 | 0.50 | p=.067 |
| JHU_APL | -0.19 | 0.36 | p<.0001 | -0.25 | 0.47 | p<.0001 | -0.28 | 0.56 | p<.0001 | -0.15 | 0.64 | p<.0001 |
| JHU_IDD | -0.49 | 0.50 | p<.0001 | -0.50 | 0.51 | p<.0001 | -0.47 | 0.51 | p<.0001 | -0.37 | 0.50 | p=.013 |
| LANL | -0.22 | 0.33 | p<.0001 | -0.26 | 0.41 | p<.0001 | -0.22 | 0.48 | p<.0001 | -0.09 | 0.52 | p=.914 |
| LNQ | -0.25 | 0.28 | p<.0001 | -0.30 | 0.37 | p=.053 | -0.23 | 0.40 | p=.005 | -0.04 | 0.45 | p<.0001 |
| UGA_CEID | -0.16 | 0.27 | p<.0001 | -0.22 | 0.38 | p=.004 | -0.18 | 0.46 | p=.017 | -0.02 | 0.53 | p=.450 |
| UpstateSU | -0.12 | 0.20 | NA | -0.14 | 0.35 | NA | -0.11 | 0.43 | NA | 0.04 | 0.52 | NA |
| UVA | 0.00 | 0.33 | p<.0001 | -0.06 | 0.40 | p<.0001 | -0.05 | 0.51 | p<.0001 | 0.10 | 0.59 | p<.0001 |

**Table 2.** Interclass correlation coefficients (ICC(2,1)) between the predicated and actual cases for all models.

| Nov 23 Forecast |  |  |  |  |
| --- | --- | --- | --- | --- |
| Team | Week 1 | Week 2 | Week 3 | Week 4 |
| Columbia | 0.97(F=67.87, p<.0001) | 0.92(F=25.60, p<.0001) | 0.82(F=10.24, p<.0001) | 0.71(F=5.95, p<.0001) |
| Ensemble | 0.98(F=84.19, p<.0001) | 0.94(F=30.15, p<.0001) | 0.83(F=10.91, p<.0001) | 0.67(F=5.13, p<.0001) |
| ISU | 0.93(F=29.80, p<.0001) | 0.80(F=9.25, p<.0001) | 0.60(F=4.01, p<.0001) | 0.42(F=2.47, p<.0001) |
| JHU_APL | 0.93(F=28.18, p<.0001) | 0.83(F=10.98, p<.0001) | 0.64(F=4.57, p<.0001) | 0.54(F=3.34, p<.0001) |
| JHU_IDD | 0.95(F=43.42, p<.0001) | 0.90(F=20.69, p<.0001) | 0.80(F=9.14, p<.0001) | 0.65(F=4.75, p<.0001) |
| LANL | 0.96(F=55.26, p<.0001) | 0.95(F=42.28, p<.0001) | 0.87(F=14.87, p<.0001) | 0.73(F=6.33, p<.0001) |
| LNQ | 0.98(F=83.62, p<.0001) | 0.96(F=45.66, p<.0001) | 0.86(F=13.58, p<.0001) | 0.76(F=7.27, p<.0001) |
| UGA_CEID | 0.97(F=64.10, p<.0001) | 0.90(F=18.33, p<.0001) | 0.74(F=6.63, p<.0001) | 0.59(F=3.91, p<.0001) |
| Upstate | 0.97(F=65.34, p<.0001) | 0.87(F=15.22, p<.0001) | 0.71(F=5.84, p<.0001) | 0.56(F=3.58, p<.0001) |
| UVA | 0.93(F=30.08, p<.0001) | 0.90(F=18.10, p<.0001) | 0.78(F=8.21, p<.0001) | 0.61(F=4.17, p<.0001) |
| Nov 30 Forecast |  |  |  |  |
| Team | Week 1 | Week 2 | Week 3 | Week 4 |
| Columbia | 0.92(F=24.08, p<.0001) | 0.73(F=6.60, p<.0001) | 0.55(F=3.49, p<.0001) | 0.55(F=3.43, p<.0001) |
| Ensemble | 0.94(F=31.66, p<.0001) | 0.81(F=9.53, p<.0001) | 0.72(F=6.21, p<.0001) | 0.74(F=6.62, p<.0001) |
| ISU | 0.92(F=25.39, p<.0001) | 0.76(F=7.22, p<.0001) | 0.58(F=3.77, p<.0001) | 0.56(F=3.52, p<.0001) |
| JHU_APL | 0.96(F=51.02, p<.0001) | 0.90(F=18.13, p<.0001) | 0.84(F=11.59, p<.0001) | 0.82(F=10.27, p<.0001) |
| JHU_IDD | 0.87(F=14.63, p<.0001) | 0.77(F=7.69, p<.0001) | 0.67(F=5.11, p<.0001) | 0.72(F=6.18, p<.0001) |
| LANL | 0.96(F=47.14, p<.0001) | 0.92(F=25.00, p<.0001) | 0.84(F=11.65, p<.0001) | 0.81(F=9.47, p<.0001) |
| LNQ | 0.93(F=30.31, p<.0001) | 0.79(F=8.77, p<.0001) | 0.66(F=4.85, p<.0001) | 0.69(F=5.51, p<.0001) |
| UGA_CEID | 0.93(F=28.56, p<.0001) | 0.79(F=8.59, p<.0001) | 0.64(F=4.57, p<.0001) | 0.64(F=4.61, p<.0001) |
| Upstate | 0.96(F=53.56, p<.0001) | 0.79(F=8.78, p<.0001) | 0.58(F=3.74, p<.0001) | 0.50(F=3.02, p<.0001) |
| UVA | 0.84(F=11.33, p<.0001) | 0.80(F=9.06, p<.0001) | 0.81(F=9.56, p<.0001) | 0.87(F=14.62, p<.0001) |

\*The ICC(2,1) reported in this table are only for counties that are present in all team/models at each forecast, as those in Table 1.

Comparison with nine CDC models showed that overall, our model yielded comparable or better accuracies than most models including the Ensemble model, particularly for short-term forecasts.

## Discussion

Realtime, accurate, and robust forecasting for the virus spread within the local government jurisdictions such as US counties is crucial for decision making. To contribute the CDC’s effort of generating an accurate and aggregated ensemble forecasting, we developed a multi-target Seq2Seq model to forecast several important indictors of COVID-19’s infection spread for the next 30 days. On average, our model generated accurate forecasts of daily incident cases, deaths, and the reproductive R numbers for the US counties.

Our model was better for predicting incident cases than for predicting incident deaths and R values. Our county-level cases forecast has been submitted and included in the CDC’s ensemble model since January 4, 2021. Although the accuracies for forecasted deaths and R values are lower than for cases, these forecasts were in highly significant agreement with the actual reported numbers as the interclass correlation results showed. Ensemble model has been used as an effective method of leveraging the power from each individual independent model and reducing the overall uncertainty (Reich, McGowan et al. 2019, Ray, Wattanachit et al. 2020). However, there is currently no coordinated efforts for creating an ensemble for county-level forecasts of death. No other models that attempted to forecast R values. Our model is one of the first to provide the real-time and accurate forecast of the county-level death and the first to forecast R values.

Uncertainty in forecasting models is common as had been observed in influenza epidemic and other serious outbreaks such as the Ebola, and Zika viruses (Eisenberg, Eisenberg et al. 2015, Moran, Fairchild et al. 2016, Kobres, Chretien et al. 2019, Zimmer, Leuba et al. 2019). Many complex and unknown features contribute to virus spread, disease severity and mortality. It is also common to have many irregularities and unforeseen discrepancies in the reporting process and database curation, which adds additional noise to the data and poses challenges for accurate forecasting. While the factors that contributed to our forecast errors and uncertainties remain to be clarified, including other local parameters, such as hospitalization and ICU saturation rates, and data from local nursing homes, may help to improve forecast accuracy for incident deaths. However, these data are not readily available for many counties. The COVID-19 pandemic continues in the US and throughout the world with substantial uncertainty in the next months due to the appearances of new strains and the logistics of vaccinating entire populations. Having data about these variables at the county level could improve forecasts in the future.

One important limitation to consider when interpretating the forecasting results is that the attempt to calculate pandemic projections, such as ours and others, was based upon only the observation of emergent cases. However, case reporting is not uniform across entities. In the US, much of the information about cases is collected and collated at the level of the county or large municipalities, which then report to a state department of health, and which in turn report to national repositories. One entity may put a strong emphasis on testing individuals who present with symptoms, whereas another may have implemented a widespread asymptomatic surveillance policy. How and which cases are identified can be dramatically affected by such policy differences and testing strategies, which were largely influenced by non-objective policy decisions and human interpretation during the course of the pandemic. In short, our machine learning model, as well as most other forecasting models of COVID-19, only learns to predict the reported cases (or deaths and other indices), which were likely biased with non-objective influences that were not uniform across reporting entities.

Since January 4, 2021, we have updated our forecast of deaths and R numbers in our Github repository each Monday (https://ylzhang29.github.io/UpstateSU-GRU-Covid/). Visualization of several useful metrics that are derived from our forecasts are provided on the GitHub page to facilitate understanding. These visualizations include projections of cases and deaths for all counties that had forecasts, and US maps of all counties to show the percentage of change in deaths and R values, and weekly cases and deaths per 100,000 population in future weeks. The code for our model’s deep learning algorithm is also available in our repository.

## Disclosure

In the past year, Dr. Faraone received income, potential income, travel expenses continuing education support and/or research support from Takeda, OnDosis, Tris, Otsuka, Arbor, Ironshore, Rhodes, Akili Interactive Labs, Sunovion, Supernus and Genomind. With his institution, he has US patent US20130217707 A1 for the use of sodium-hydrogen exchange inhibitors in the treatment of ADHD. He also receives royalties from books published by Guilford Press: Straight Talk about Your Child’s Mental Health, Oxford University Press: Schizophrenia: The Facts and Elsevier: ADHD: Non-Pharmacologic Interventions. He is Program Director of www.adhdinadults.com.

## Data Availability

The data used in the forecasting models are available from public repositories and all forecast results are available in our github repository, and the covid19forecasthub.org

https://github.com/ylzhang29/UpstateSU-GRU-Covid

https://github.com/reichlab/covid19-forecast-hub/tree/master/data-processed/UpstateSU-GRU

## Acknowledgement

Dr. Zhang-James is supported by the European Union’s Seventh Framework Programme for research, technological development and demonstration under grant agreement no 602805 and the European Union’s Horizon 2020 research and innovation programme under grant agreements No 667302.

Dr. Faraone is supported by the European Union’s Seventh Framework Programme for research, technological development and demonstration under grant agreement no 602805, the European Union’s Horizon 2020 research and innovation programme under grant agreements No 667302 & 728018 and NIMH grants 5R01MH101519 and U01 MH109536-01.

---

**Supplementary Figure 1.**
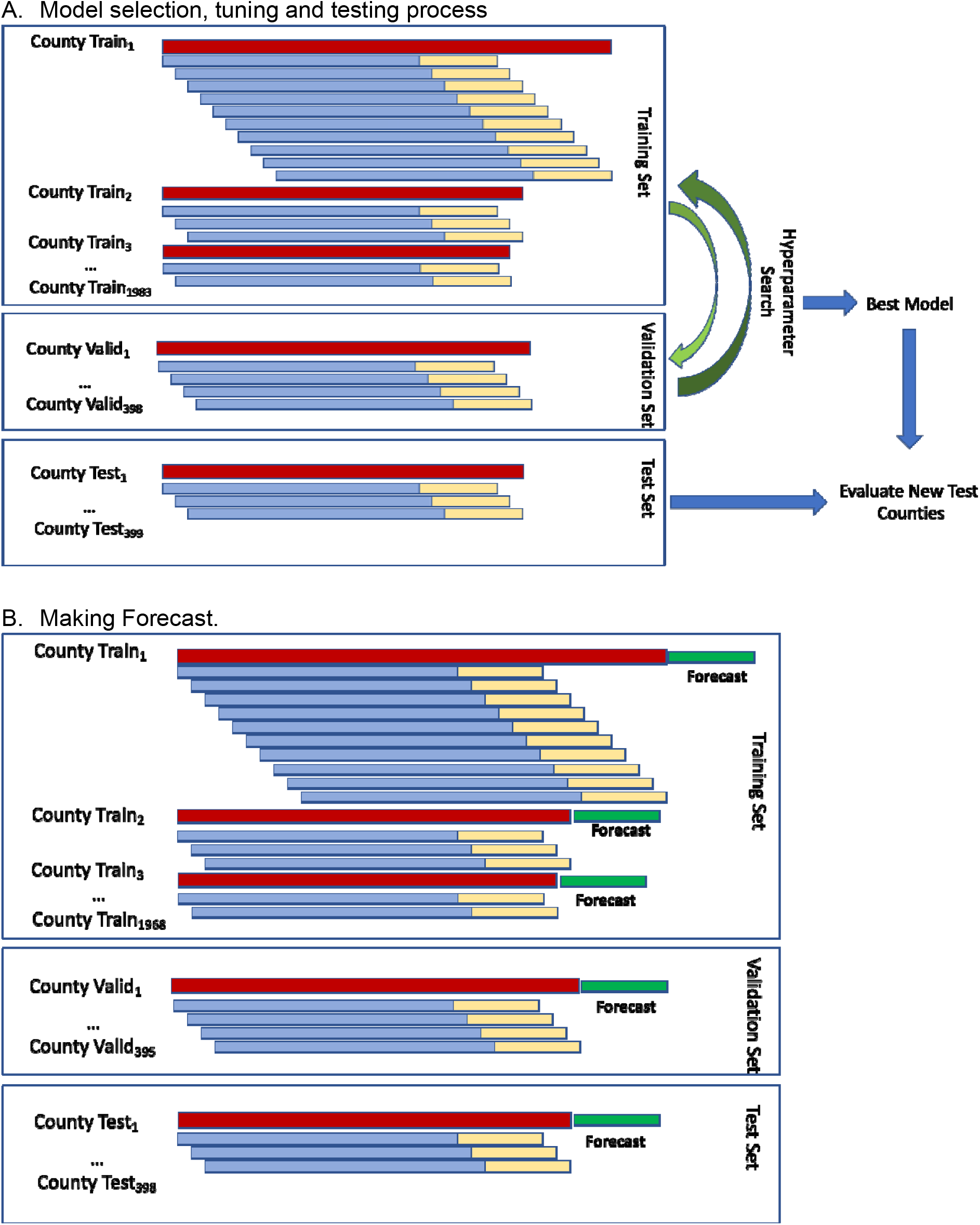
ML setup.

**Supplementary Figure 2.**
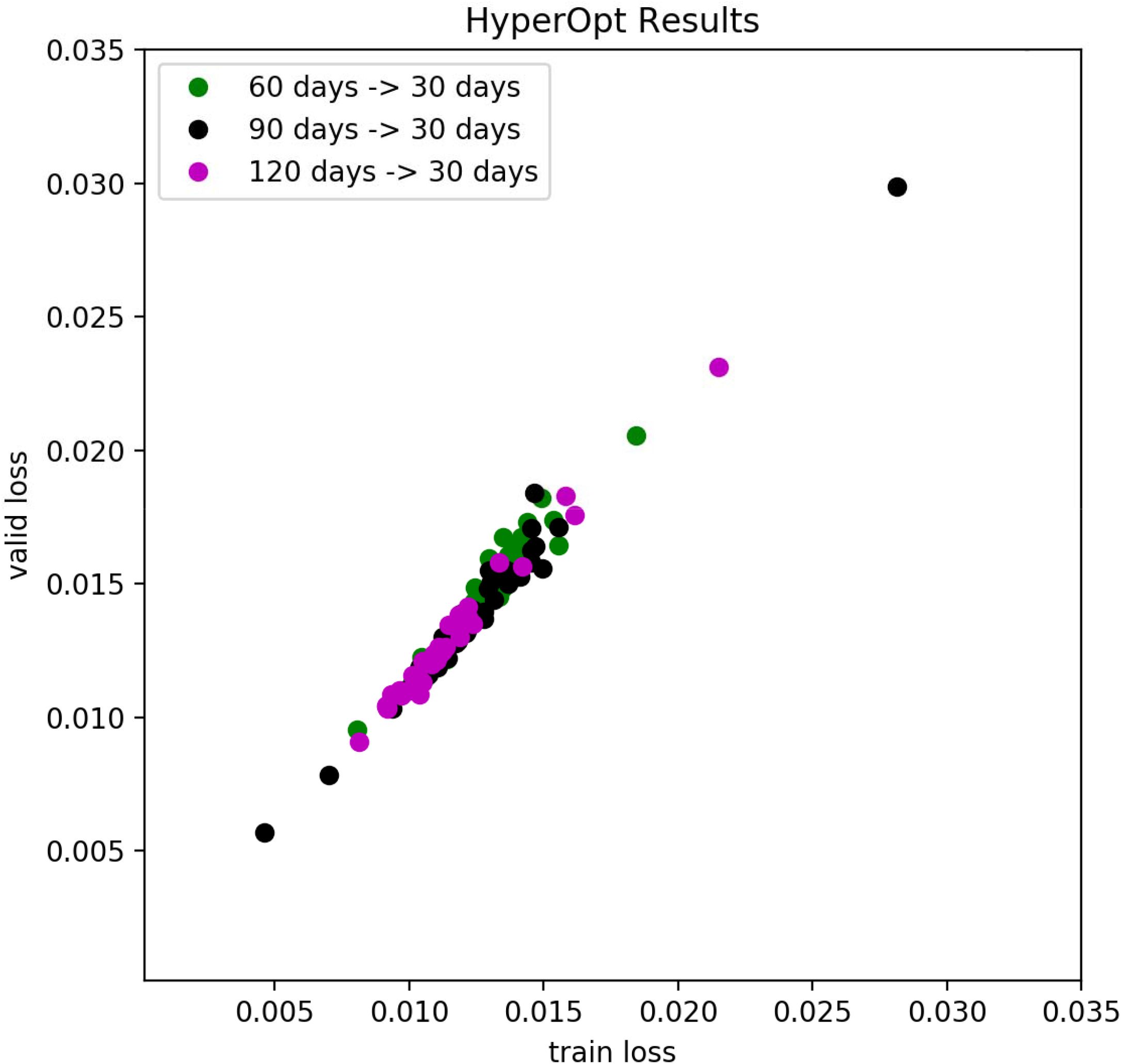
HyperOpt results of various input lenghth.

**Supplementary Figure 3.**
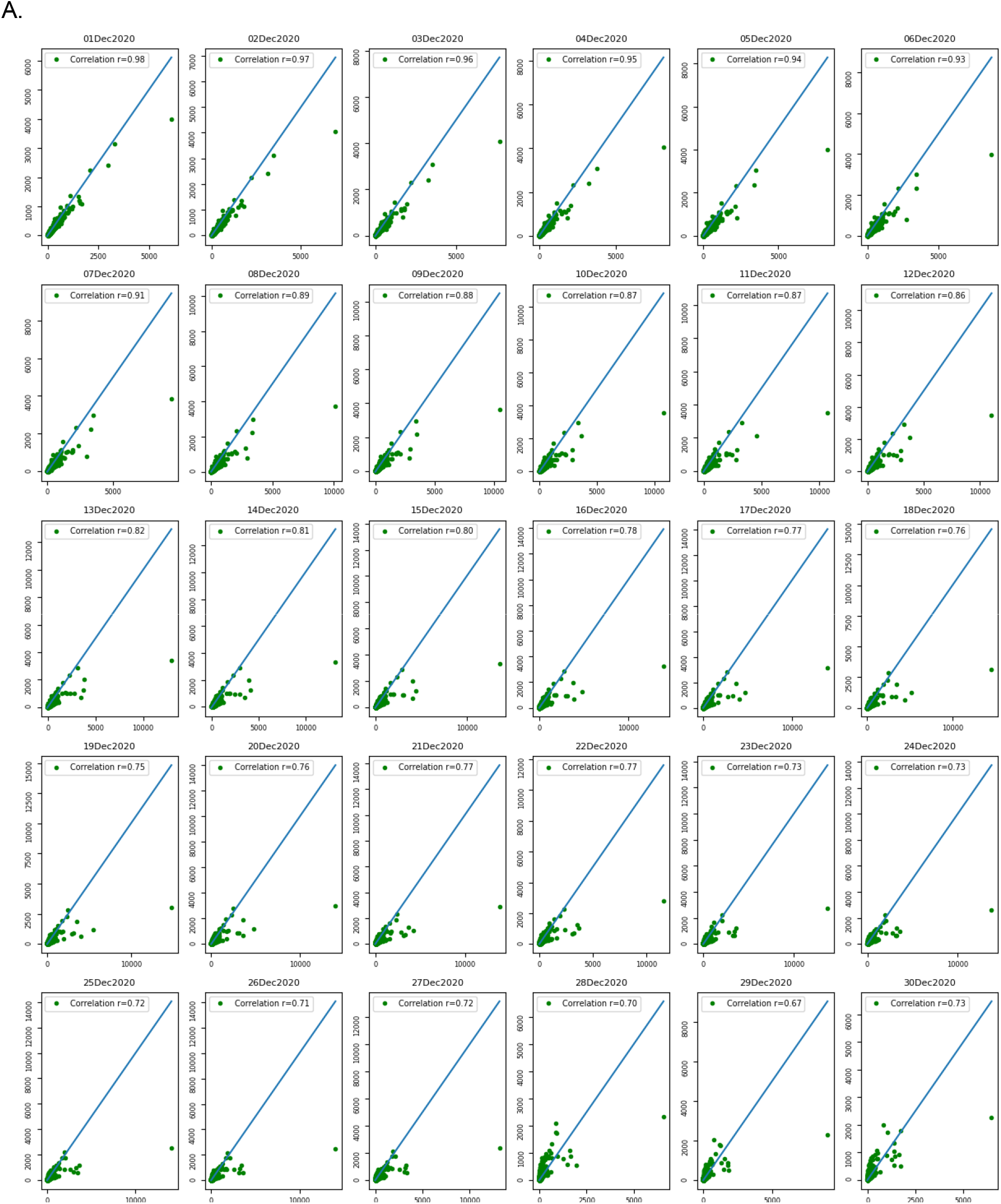
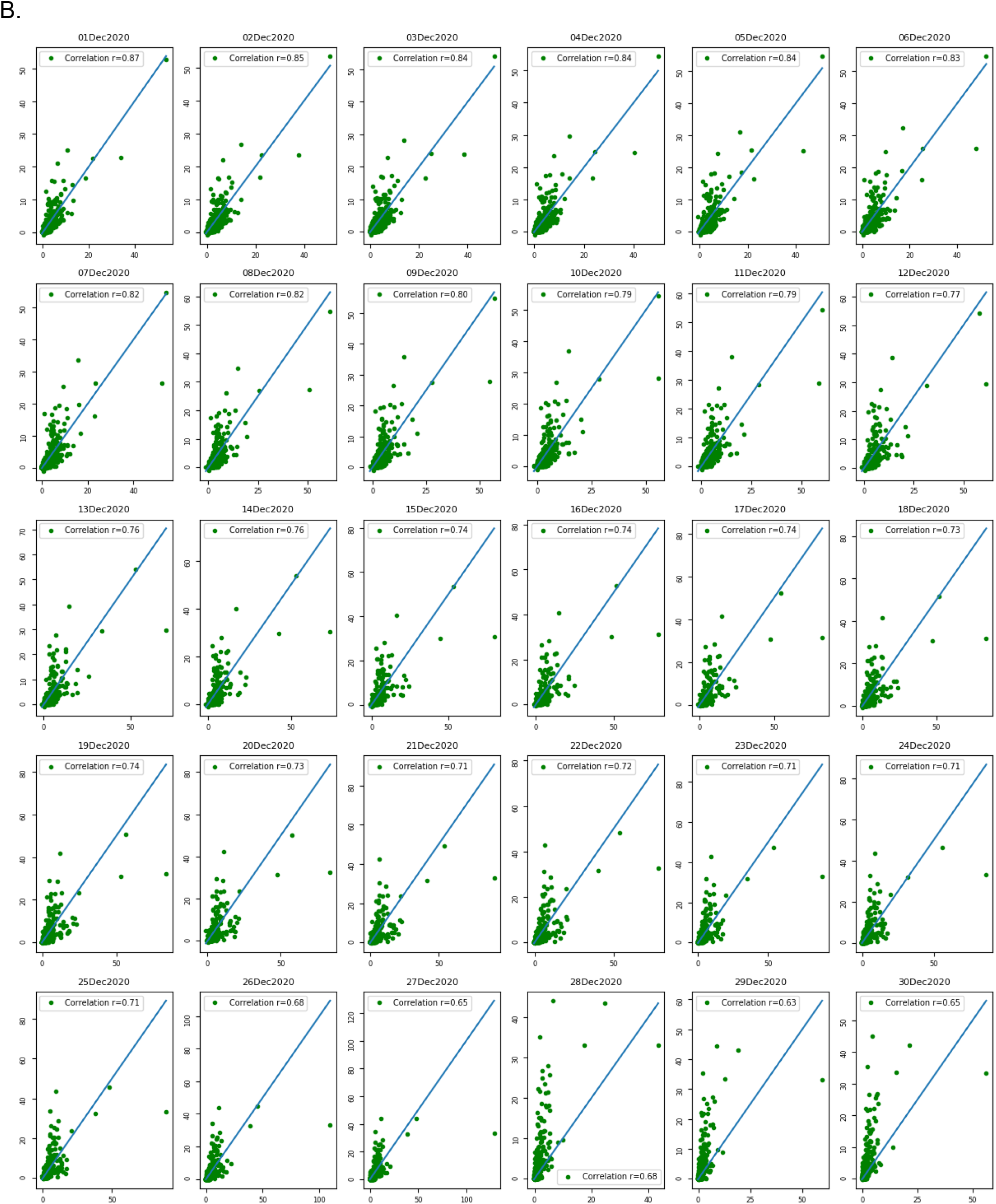
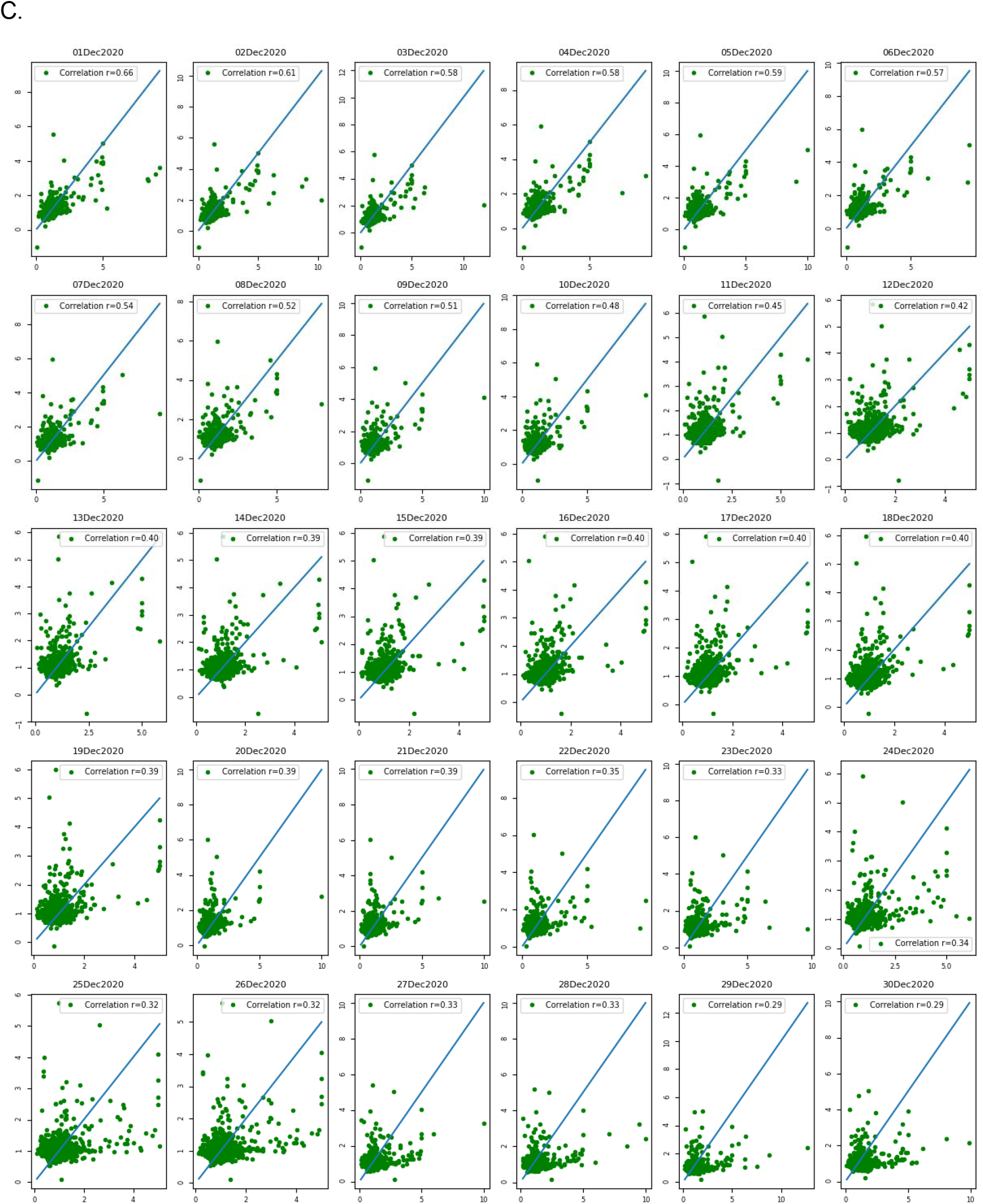
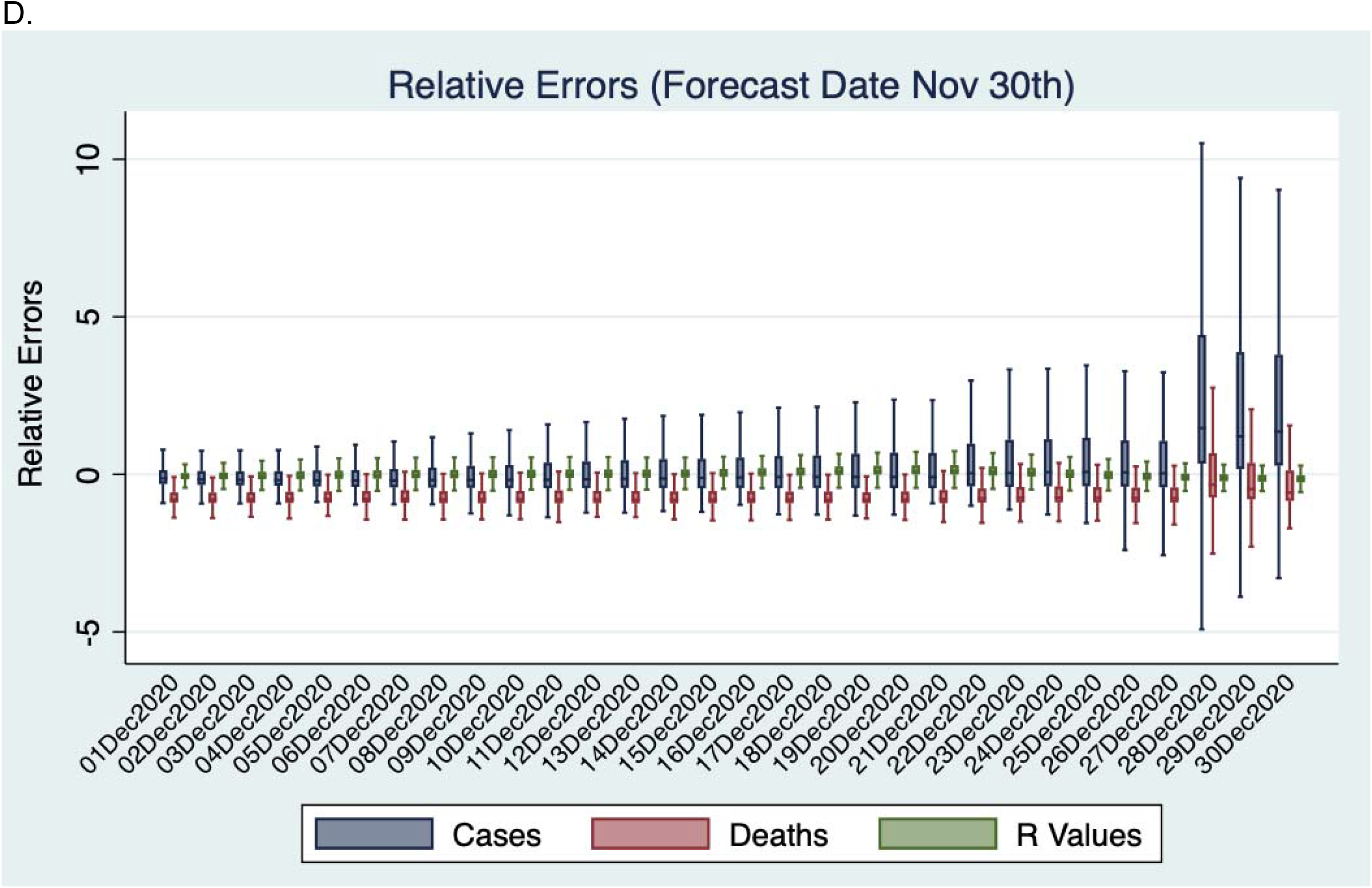
**A**. Daily predicted cases (y axis) vs actual cases (x axis) over the next 30 days from the forecast date Nov 30^th^, 2020. **B**. Daily predicted deaths (y axis) vs actual deaths (x axis) over the 30 days forecast. **C**. Daily predicted R values (y axis) vs R calculated from the actual cases (x axis) over the 30 days forecast. **D**. Relative Error distributions.

**Supplementary Table 1.**
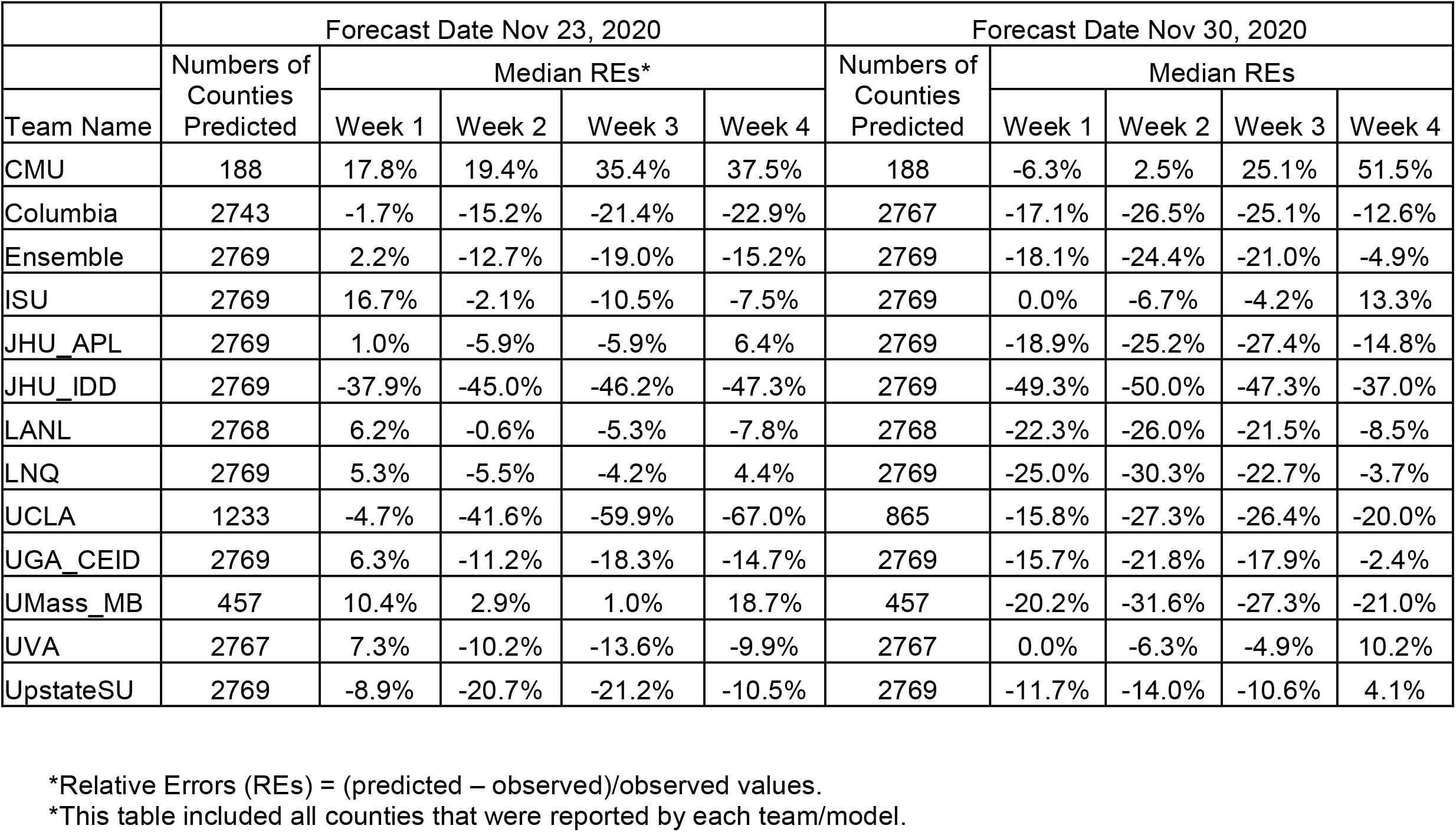
Median relative errors for county-level case forecast from different teams.

## References

Bergstra, J., D. Yamins and D. D. Cox (2013). Making a Scienceof Model Search: Hyperparameter Optimizationin Hundredsof Dimensions for Vision Architectures.. The 30thInternational Conference on Machine Learning (ICML 2013), Atlanta,Gerorgia,, JMLR Workshop and ConferenceProceedings.

Cori, A., N. M. Ferguson, C. Fraser and S. Cauchemez (2013). “A New Framework and Software to Estimate Time-Varying Reproduction Numbers During Epidemics.” American Journal of Epidemiology 178(9): 1505–1512.

Dey, R. and F. M. Salem (2017). Gate-variants of Gated Recurrent Unit (GRU) neural networks. 2017 IEEE 60th International Midwest Symposium on Circuits and Systems (MWSCAS).

Dong, E., H. Du and L. Gardner (2020). “An interactive web-based dashboard to track COVID-19 in real time.” The Lancet Infectious Diseases 20(5): 533–534.

Eisenberg, M., J. Eisenberg, J. P. D’Silva, E. Wells, S. Cherng, Y.-H. Kao and R. Meza (2015). “Forecasting and Uncertainty in Modeling the 2014-2015 Ebola Epidemic in West Africa.” arXiv: Populations and Evolution.

Kobres, P. Y., J. P. Chretien, M. A. Johansson, J. J. Morgan, P. Y. Whung, H. Mukundan, S. Y. Del Valle, B. M. Forshey, T. M. Quandelacy, M. Biggerstaff, C. Viboud and S. Pollett (2019). “A systematic review and evaluation of Zika virus forecasting and prediction research during a public health emergency of international concern.” PLoS Negl Trop Dis 13(10): e0007451.

Moran, K. R., G. Fairchild, N. Generous, K. Hickmann, D. Osthus, R. Priedhorsky, J. Hyman and S. Y. Del Valle (2016). “Epidemic Forecasting is Messier Than Weather Forecasting: The Role of Human Behavior and Internet Data Streams in Epidemic Forecast.” J Infect Dis 214(Suppl_4): S404–s408.

Ray, E. L., N. Wattanachit, J. Niemi, A. H. Kanji, K. House, E. Y. Cramer, J. Bracher, A. Zheng, T. K. Yamana, X. Xiong, S. Woody, Y. Wang, L. Wang, R. L. Walraven, V. Tomar, K. Sherratt, D. Sheldon, R. C. Reiner, B. A. Prakash, D. Osthus, M. L. Li, E. C. Lee, U. Koyluoglu, P. Keskinocak, Y. Gu, Q. Gu, G. E. George, G. España, S. Corsetti, J. Chhatwal, S. Cavany, H. Biegel, M. Ben-Nun, J. Walker, R. Slayton, V. Lopez, M. Biggerstaff, M. A. Johansson and N. G. Reich (2020). “Ensemble Forecasts of Coronavirus Disease 2019 (COVID-19) in the U.S.” medRxiv: 2020.2008.2019.20177493.

Reich, N. G., C. J. McGowan, T. K. Yamana, A. Tushar, E. L. Ray, D. Osthus, S. Kandula, L. C. Brooks, W. Crawford-Crudell, G. C. Gibson, E. Moore, R. Silva, M. Biggerstaff, M. A. Johansson, R. Rosenfeld and J. Shaman (2019). “Accuracy of real-time multi-model ensemble forecasts for seasonal influenza in the U.S.” PLOS Computational Biology 15(11): e1007486.

Zimmer, C., S. I. Leuba, T. Cohen and R. Yaesoubi (2019). “Accurate quantification of uncertainty in epidemic parameter estimates and predictions using stochastic compartmental models.” Statistical methods in medical research 28(12): 3591–3608.

